# Estimating the instantaneous reproduction number (*R*_*t*_) *by using particle filter*

**DOI:** 10.1101/2023.07.09.23292422

**Authors:** Yong Sul Won, Woo-Sik Son, Sunhwa Choi, Jong-Hoon Kim

## Abstract

**Background:** Monitoring the transmission of coronavirus disease 2019 (COVID-19) requires accurate estimation of the effective reproduction number (*R*_*t*_). However, existing methods for calculating *R*_*t*_ may yield biased estimates if important real-world factors, such as delays in confirmation, pre-symptomatic transmissions, or imperfect data observation, are not considered.

**Method:** To include real-world factors, we expanded the susceptible-exposed-infectious-recovered (SEIR) model by incorporating pre-symptomatic (P) and asymptomatic (A) states, creating the SEPIAR model. By utilizing both stochastic and deterministic versions of the model, and incorporating predetermined time series of *R*_*t*_, we generated simulated datasets that simulate real-world challenges in estimating *R*_*t*_. We then compared the performance of our proposed particle filtering method for estimating *R*_*t*_ with the existing EpiEstim approach based on renewal equations.

**Results:** The particle filtering method accurately estimated *R*_*t*_ even in the presence of data with delays, pre-symptomatic transmission, and imperfect observation. When evaluating via the root mean square error (RMSE) metric, the performance of the particle filtering method was better in general and was comparable to the EpiEstim approach if perfectly deconvolved infection time series were provided, and substantially better when *R*_*t*_ exhibited short-term fluctuations and the data was right truncated.

**Conclusions:** The SEPIAR model, in conjunction with the particle filtering method, offers a reliable tool for predicting the transmission trend of COVID-19 and assessing the impact of intervention strategies. This approach enables enhanced monitoring of COVID-19 transmission and can inform public health policies aimed at controlling the spread of the disease.

## 1. Introduction

Since its first identification in Wuhan, China in December 2019 (Lu et al., 2020; Phelan et al., 2020), the coronavirus disease-19 (COVID-19) has spread worldwide, and the characteristics of disease dynamics have changed considerably over the two-year-long pandemic. To prevent further infections, many countries have implemented various interventions, such as social distancing, contact tracing, border closures, and vaccinations. These interventions have motivated the development of quantitative techniques that provide policymakers with the ability to monitor changes in disease transmission over time and evaluate the performance of intervention programs in near real-time.

One of the key metrics used to monitor disease transmission is the effective reproduction number (*R*_*t*_), which is the average number of new infections caused by an infectious individual in a population consisting of both susceptible and non-susceptible hosts. There are two main approaches for estimating *R*_*t*_ from case incidence data. The first approach is to treat the cases occurring at time t as primary cases and calculate onward secondary transmissions from the primary cases. This approach, known as the case reproduction number, was first introduced in 2004 (Wallinga & Teunis, 2004). The second approach treats the cases occurring at time t as secondary cases reproduced by cases occurring prior to time t, with weights determined by the generation interval distribution. The estimates inferred from this latter approach are called instantaneous reproduction number *R*_*t*_ (Fraser, 2007) and have become more widely adopted following a study in the field (Cori et al., 2013). The instantaneous reproduction number *R*_*t*_ is considered a better indicator for ongoing dynamics of transmission (Gostic et al., 2020).

Although calculating *R*_*t*_ is straightforward (Cori et al., 2013; Cori, 2021), applying the method mechanically without considering the context of the data can lead to a biased estimate (Gostic et al., 2020). For example, one real-world context is delay from infection to symptom onset or confirmation. While ideally *R*_*t*_ calculation needs to be based on the time series of infections to give timely estimates, most COVID-19 data are given as time series of case confirmation or death. If the method is simply applied to the dataset with delay (e.g., time series of case confirmation), the inferred *R*_*t*_ will on average indicate the reproduction number of the past (e.g., *t* – average delay from infection to confirmation). The paper by Gostic et al. (Gostic et al., 2020) describes a way to overcome this challenge by deconvolving confirmation time series to infer infection time series and then applying the method to the inferred infection time series. There are other real-world contexts such as uncertainty in parameter estimates (e.g., generation interval), imperfect observation, or right truncation as described in the previous study. Another real-world context omitted in previous study (Gostic et al., 2020) is the generation interval of COVID-19, which is the interval between infection times of successive cases (Fine, 2003). Since COVID-19 can be transmitted during the incubation period before symptoms appear, the serial interval (i.e., the interval between symptom onsets of successive cases) (Porta, 2014) may not be a good substitute for the generation interval (Ganyani et al., 2020).

To address this issue, we propose an estimation strategy for instantaneous *R*_*t*_ using a particle filtering (also known as sequential Monte Carlo) method (Arulampalam et al., 2002). PF is a popular choice for calibrating nonlinear dynamical systems such as compartment models of infectious disease transmission in epidemiology (Yang et al., 2014; Dukic et al., 2012; Calvetti et al., 2021; Safarishahrbijari et al., 2021). We explored the potential to use a dynamic compartmental model of disease transmission (e.g., SEIR model) in which the transmission rate parameter is estimated using a PF method. By using a dynamic model, researchers can better address the previously mentioned issues (i.e., delay from infection to confirmation and the correct use of generation interval) and create a framework to test hypotheses or potential impact of intervention programs. While there are several successful data assimilation models (e.g., Kalman filter, particle filter, ensemble Kalman filter) to estimate latent variables or parameters and *R*_*t*_ (Yang et al., 2014; Kucharski et al., 2020; Arroyo-Marioli et al., 2021), this work is the first to systematically investigate the PF using simulated data in the context of *R*_*t*_.

## 2. Materials and Methods

### 2.1. Compartment model (SEPIAR model)

To capture realistic *R*_*t*_ of COVID-19, we adapt the well-known SEIR modeling approach and introduce the SEPAIR model equations as follows:

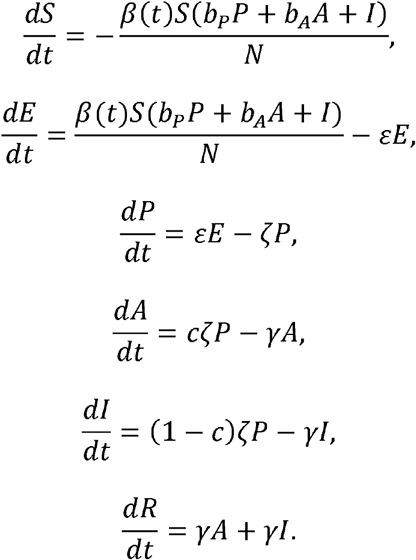

Here, as depicted in Figure 1, the population is divided into 6 groups, including susceptible (S), exposed (E), pre-symptomatic infectious (P), symptomatic infectious (I), asymptomatic infectious (A), and removed (R) (i.e., isolated, recovered, or otherwise no longer infectious) so that the total population (*N*) remains constant for all times t (in days), i.e., N= S + + P+ + A + R. In addition, the SEPIAR model accounts for infections during latency and asymptomatic infections in the following sense: *β* (t) is the transmission rate at time t that susceptible hosts become exposed in contacts with the hosts in all three *P, I* and *A* groups, where the scaling factors (*b*_*p*_ and *b*_*A*_), each ranging between 0 and 1, apply for the transmissibility whilst at the stages P and A respectively compared to the stage. We took bp = bA =1 for simplicity in this study as the primary scenario; the mean residence time in the stage P (1/ *ζ*) is fixed at 2.5 days, which is calculated as the difference between the mean incubation period (1/ *η* =5 days) and the mean latent period (1/ ε = 2.5 days); c is the probability of entering the stage A on leaving the stage P and we set that c= 0.3 in this study; and finally the delay from onset to isolation (1/ *γ*) is assumed to be 2.5 days. The values of the parameters are summarized in Table 1.

**Fig. 1.**
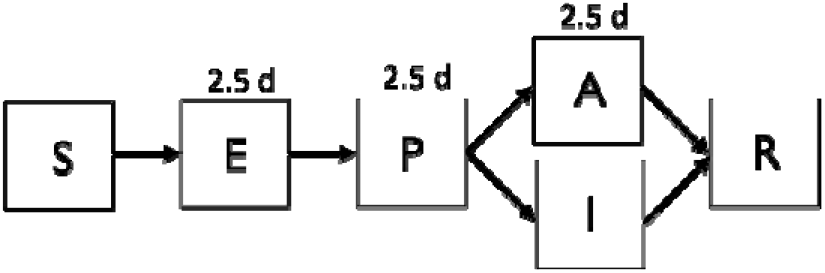
Compartmental Flow of the SEPIAR model.

**Table 1.** Summary of Model Evaluation.

|  | Types of uncertainty | Data type | Dataset |
| --- | --- | --- | --- |
| **Test 1** | No delays; Deterministic model | Daily infection | $\mathcal{D}_1$ |
| **Test 2** | Reporting delay; Deterministic model | Daily confirmation | $\mathcal{D}_1$ |
| **Test 3** | Reporting delay; Stochastic model | Daily confirmation | $\mathcal{D}_2$ |

#### 2.1.1. Effective reproduction number

We define the effective reproduction number, *R*_*t*_ (*t*), by the product of time-varying transmission rate and infectious period and, for the SEPAIR model, *R*_*t*_ (t) can be expressed as:

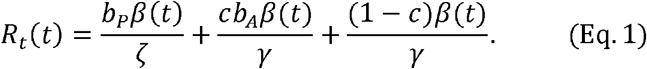

The above formulation of *R*_*t*_ (t) will be used for the rest of this paper.

### 2.2. Data sets

We use simulated data by the model in which we know the governing parameter values. The simulated data consists of the time series of daily infection, symptom onset, confirmation predicted by the SEPIAR model with a pre-determined trajectory (Figure 2a and 2b). The pre-determined trajectory was designed to be simple but still to capture the reality of SARS-CoV 2 transmission that would hover around the threshold value of one because of human interventions such as social distancing or mask wearing (Figure 2a). We included various real-world aspects of data collection during a COVID epidemic by adding uncertainties to the model in a stepwise manner. As the results, we generated two sets of simulated data (𝒟_1_𝒟_2_) as below.

**Fig. 2.**
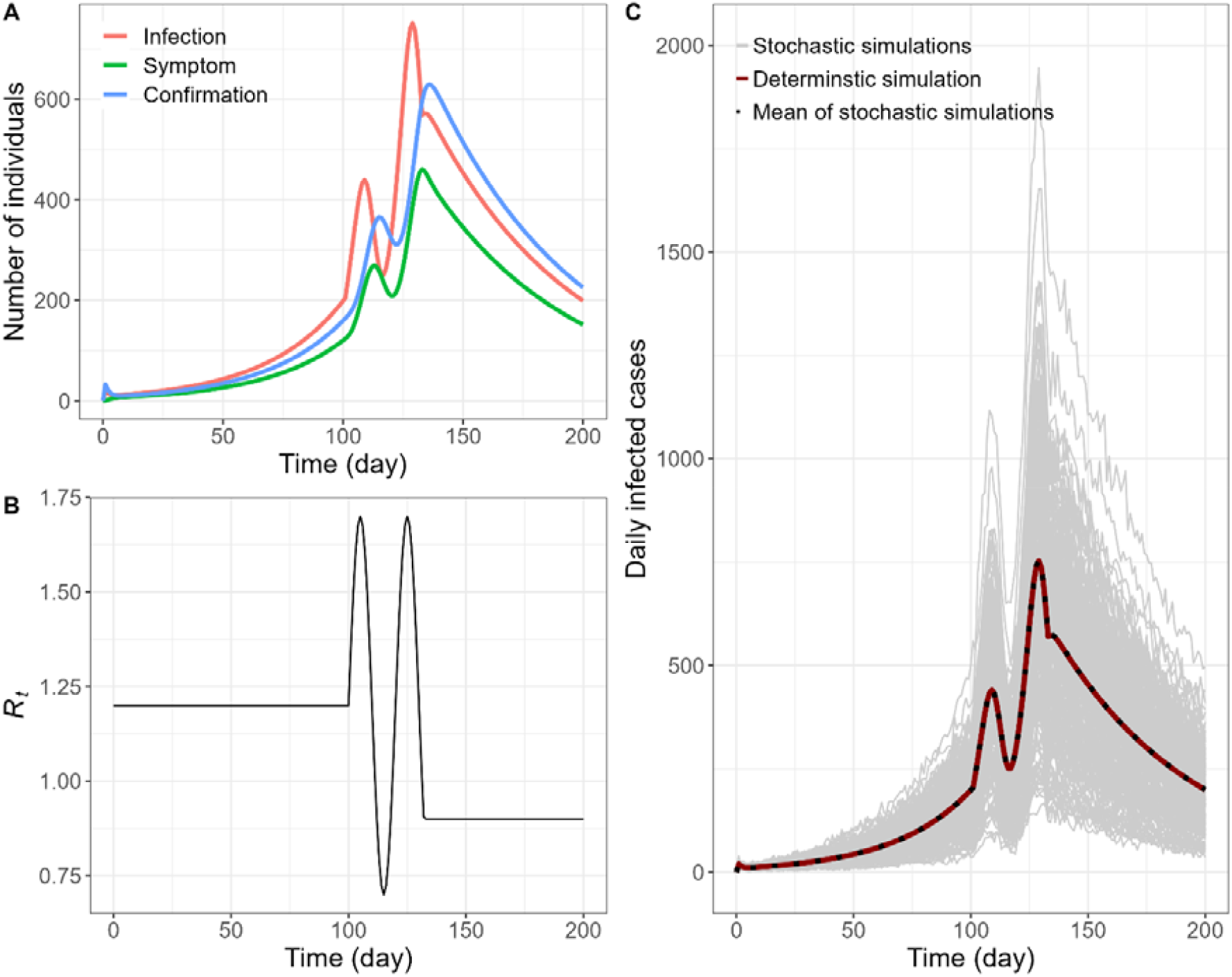
Simulations of SEPAIR model. A. Time series of infection (red), symptom (green), and confirmation (blue) based on deterministic simulations of SEPIAR model. **B**. Pre-determined daily effective reproduction number, R_t_. C. Time series of confirmation based on 2,000 stochastic simulations of SEPIAR model (grey), deterministic simulation (red), and the mean of the stochastic simulations (dotted).

i. The simulated data 𝒟_1_is simply obtained by solving a deterministic model (SEPIAR equations) under the condition of perfect observation (i.e., asymptomatic as well as symptomatic cases are all confirmed). We track cumulative sum of the states (e.g., *E, P, I*) over the simulation period and then calculate increments for the daily size of infections, symptoms, and confirmations, respectively. Here, the daily infection time series represent the exact case counts at the time of infection, whereas the daily confirmation series act as the case counts with reporting delay.
ii. The simulated data 𝒟_2_is obtained by solving a stochastic model of SEPAIR by Gillespie’s direct method (Gillespie, 1976; Gillespie, 1977). The assumption of perfect observation remains valid here. Averages of large samples would retrieve the cases of 𝒟 1 (Figure 2c).

All the data generations were implemented in programming language R and the differential equations were solved using the deSolve package. The codes used to generate the simulated data sets and the analyses conducted in this paper are available on the GitHub page of the last author (https://github.com/kimfinale/pfilterCOVID).

We also included a possibility of misspecification of serial interval when using the EpiEstim method to estimate *R*_*t*_for a COVID-19 outbreak. Serial interval is a critical input for the EpiEstim method as a proxy for the generation interval. For COVID-19, pre-symptomatic transmissions may lead that the serial interval is a poor proxy for the generation interval. Estimated mean serial intervals based on the field data are approximately 5 days (Alene et al., 2021; Rai et al., 2021; Nishiura et al., 2020; Linton et al., 2020) which are close to or shorter than the incubation period (Linton et al., 2020). Our simulated data sets were generated under the assumption that the mean incubation period is 5 days and the mean generation time is 6.25 days. To evaluate the impact of misspecification of serial interval, we set the serial interval to be 5 days as an input to the EpiEstim method and examined how estimated *R*_*t*_ ‘s are influenced by this misspecification.

### 2.3. Model evaluation with simulated data

How well the PF can recover the true value of can be assessed by computing the root mean squared error (RMSE) defined as

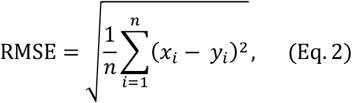

where *x*_*i*_is a true (or observed) value and *y*_*i*_is an estimate of*x*_*i*_ for a total estimation size of. The simulated datasets (𝒟_1_𝒟_2_) were used to test systematically the robustness of our model under various types of noises and delays. We first estimated by *R*_*t*_using the time series of daily infection with perfect observation. This may represent an ideal data set which we would not be able to obtain in reality, but serves as the baseline on which our method should work. Then, the estimation model is applied to the time series that contains delays (i.e., the daily confirmation) and stochasticity. We note that individual outputs from a stochastic model vary by simulation and therefore may be regarded as imperfect observation. These can be summarized as three tests as illustrated below in Table 1. We assume all infected people are detected (i.e., complete observation) in the simulated data sets as this assumption makes our analyses simple. However, the data set includes stochastic time series, which may account for varying probability detection of over the time series.

We also use these tests to compare the performance of our model against the benchmarking method, EpiEstim (Cori, 2021) and deconvolution. To infer the time series of infection by deconvolving the time series of confirmation, we need information on the delay from infection to confirmation (i.e., from to R state). As shown in Figure 1, it equals the sum of three independent exponential distributions, Exp(λ = 1/2.5). According to Exp(*λ*) = Γ(α = 1, β = *λ*) and Γ(al,β) + Γ(a2, β) = Γ(al + a2, β), the time distribution from infection to confirmation can be represented by the gamma distribution Γ(3,1/2.5). Then, to estimate *R*_*t*_using EpiEstim from the inferred time series of infection, the distribution of serial interval should be specified. The serial interval represents the time between the clinical onsets of successive cases (Fine, 2003) and it is naturally expressed as the sum of the incubation period and disease age at transmission (Nishiura, 2009). The distribution of serial interval satisfies the following convolution 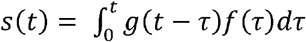, where 9 is the distribution of disease age and f is the distribution of incubation period. As a result, the mean and standard deviation of the serial interval are 6.247 and 4.138, respectively.

### 2.4. Particle Filtering Method

The particle filtering method is employed to estimate the distribution of effective reproduction number *R*_*t*_by using the definition (Eq.1). Let us write *x*_0:t_ = *x*_0_,…, *x*_*t*_ and *Y*_l:t_ = *Y*_l_,…,*Y*_t_ to denote the vector of latent variables and observation up to time t, respectively. The choice of state vector and observation are as follows:

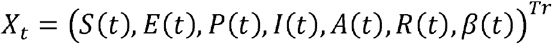

and *Y*_l:t_ = times series of confirmation up to time t, respectively. The time-varying transmission rate β (t) was assumed to follow a geometric Brownian motion (GBM) (Pavliotis, 2014) satisfying the stochastic differential equation: *d* log β (t) = *σ dw*_*t*_, where *w*_*t*_ is a Brownian motion and a is a constant diffusion constant. Together with the SEPIAR equations above, it gives rise to a set of stochastic differential equations whose solutions is x_t_. The distribution of *R*_*t*_can be obtained directly from the definition (Eq.1) once the distribution of *β* (t) is inferred by particle filter.

#### 2.4.1. Ideas of Particle Filter

Latent variables, {*X*_t_}_t ε ℕ_, are modeled as a Markov process with an initial distribution p(x0) and transition probabilities p(x_t_|x_t-l_), t ≥ 1. The observations, {*X*_t_}_t ε ℕ_are assumed to be conditionally independent of the process {*X*_t_}_t ε ℕ_ and of the likelihood (marginal density) p(*Y*_t_|*X*_t_). More precisely, the particle filtering algorithm estimates the posterior distribution p(x_0:t_|Y_l:t_), or the filtering distribution p(*X*_t_|*Y*_*l:t*_) recursively for a training period, t = 1,2, …,t_*p*_, in the framework of Monte Carlo method, that is, for N -number of approximating particles 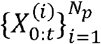

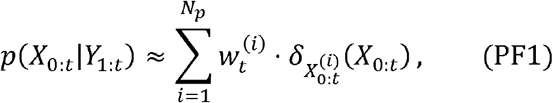

and

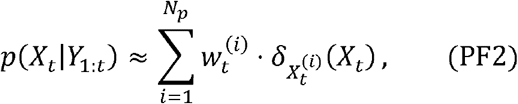

where 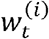 are weight values corresponding to the Dirac measures 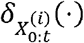 and 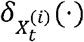

Notice that integrating the equation (PF1) with respect to *x*_*0:t-1*_ yields the formulation (PF2).

#### 2.4.2. Bootstrap Filter with Backward Selection

We implemented a bootstrap particle filter followed by a backward sampling. The modelling assumptions of SEPIAR and GBM were used when sampling from prior distributions.

i. Initialization (t= 0)
  - For = 1, …,N, sample 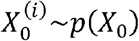.
ii. Importance sampling step (t= 1→ *t*_*p*_)
  - For = 1, …,N, sample 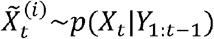 and set 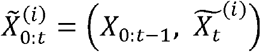.
  - For = 1, …,N, assign the importance weights proportional to the likelihood, i.e.,

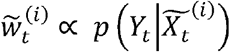

and then normalize the weights.
iii. Resampling step (t= 1 → *t*_*p*_)
  - Resample with replacement 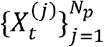 from the set 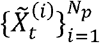 according to the normalized importance weights 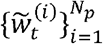 and save the resampling path as A _*t*_, i.e., (*j*)^th^ component of A_*t*_ is the index *k* such that 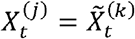
  - Assign uniform weights to the resampled particles, i.e.,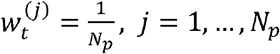
  - Set t ←t + 1 and go back to step (2) unless t= t _*p*_.
iv. Backward sampling step (t= t_*p*_ → 1)
  - Sample a particle index 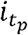 from the resampling path 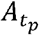 according to the importance weights 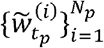
  - For t= t -1, …, 1, choose indices t as the component of A_t+l_ at (*i* _*t+l*_)^*th*^ place.
  - Select a trajectory of particles 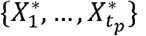 by 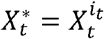 for t= 1,…, t_*p*_.
  - Iterate the above steps N_b_ -many times.

The algorithm is formed of two parts: forward filtering (step (1) – step (3)), where prediction and updating of filtering distribution takes place; and backward sampling (step (4)), where a trajectory of particles is selected backward in time on a basis of best fitting particle at the end of training period. In this study, we used 10,000 particles and 1,000 backward iterations (i.e., N = 10,000 and Nb =1,000).

## 3. Results

### 3.1. Estimation of R_*t*_ based on the particle filtering method

#### 3.1.1. Inferring R_*t*_ using infection and confirmation times series based on a deterministic simulation model

Under the ideal scenario in which we have access to the time series of daily infected cases with complete observation, the particle filtering method should be able to retrieve the pre-defined *R*_*t*_and other state variables. We first apply the PF method to the dataset (𝒟1) that was generated by a deterministic simulation of SEPAIR model. With a setting of 10,000 particles and 1,000 backward sampling (i.e., *N*_*p*_ = 10,000 and *N*_*b*_ =1,000), the resulting posterior distribution of *R*_*t*_captures true *R*_*t*_very well (Fig 3A). Given a time series of daily confirmed cases, the deviation between the true and the inferred *R*_*t*_ ‘s and the uncertainty around inferred *R*_*t*_is larger, especially in the last 8 days or so, which is similar to the delay from infection to confirmation (i.e., mean delay = 7.5 days). But the PF inferred *R*_*t*_still captures the overall shape and magnitude of the true *R*_*t*_(Fig 3B).

**Fig. 3.**
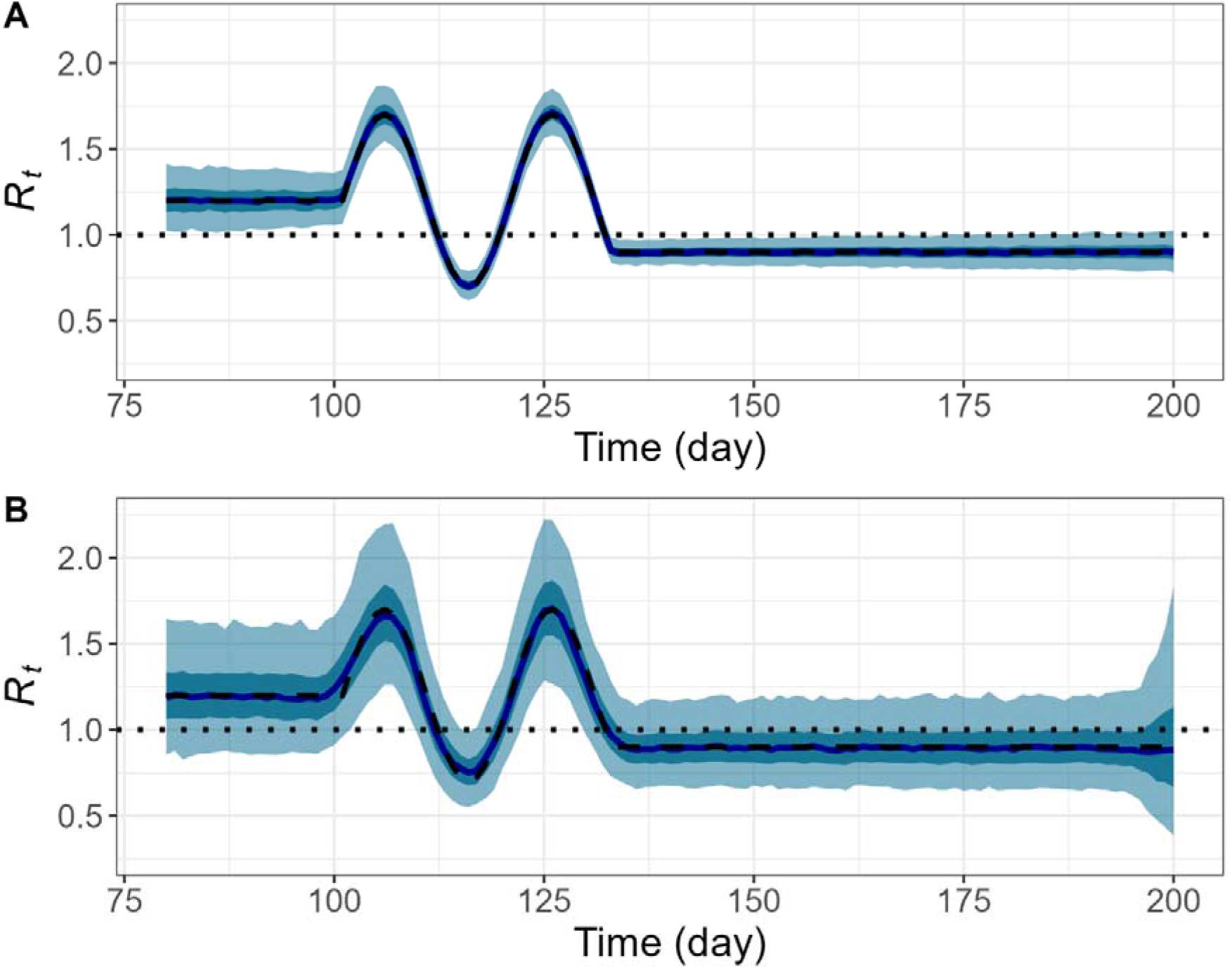
Daily reproduction number, *R*_*t*_, inferred by particle filtering applied on deterministic SEPAIR models with N = 10,000 and Nb =1,000. Pre-defined *R*_*t*_(black dashed), reference line (black dotted), median of estimated *R*_*t*_(dark blue), interquartile range of estimated *R*_*t*_(dark cyan shaded), and middle 95% of estimated *R*_*t*_(light cyan shaded). **A**. Infection time series as observation. **B**. Confirmation time series as observation.

#### 3.2.1. Inferring *R*_*t*_using confirmation times series based on a stochastic simulation model

The particle filtering method is also capable of recovering the pre-defined even though observed confirmations contain uncertainties such as reporting delay. To reflect real-world observations, three time series of daily confirmed cases were chosen on the basis of low, medium, and high incidence from stochastic simulations of the SEPAIR model, i.e., dataset

(Fig 4A). Again, with a setting of 10,000 particles and 1,000 backward sampling, the resulting posterior distributions *R*_*t*_ of replicate true*R*_*t*_ quite closely regardless of the raw intensity of observations (Fig 4B, 4C, 4D). In addition, as in the case of deterministic simulation, the range of estimates widens approximately in the last 8 days of the estimation. However, the estimations based on the greatest number of raw cases are less vulnerable to sudden changes of transmission patterns, for example the peaks on day 107 and 125 (Fig 4B), compared to over- or under-estimation of *R*_*t*_(Fig 4C, 4D). Those peaks correspond to the beginning of a new phase of transmission, each leading to a surge in daily confirmed cases (Fig 4A).

**Fig. 4.**
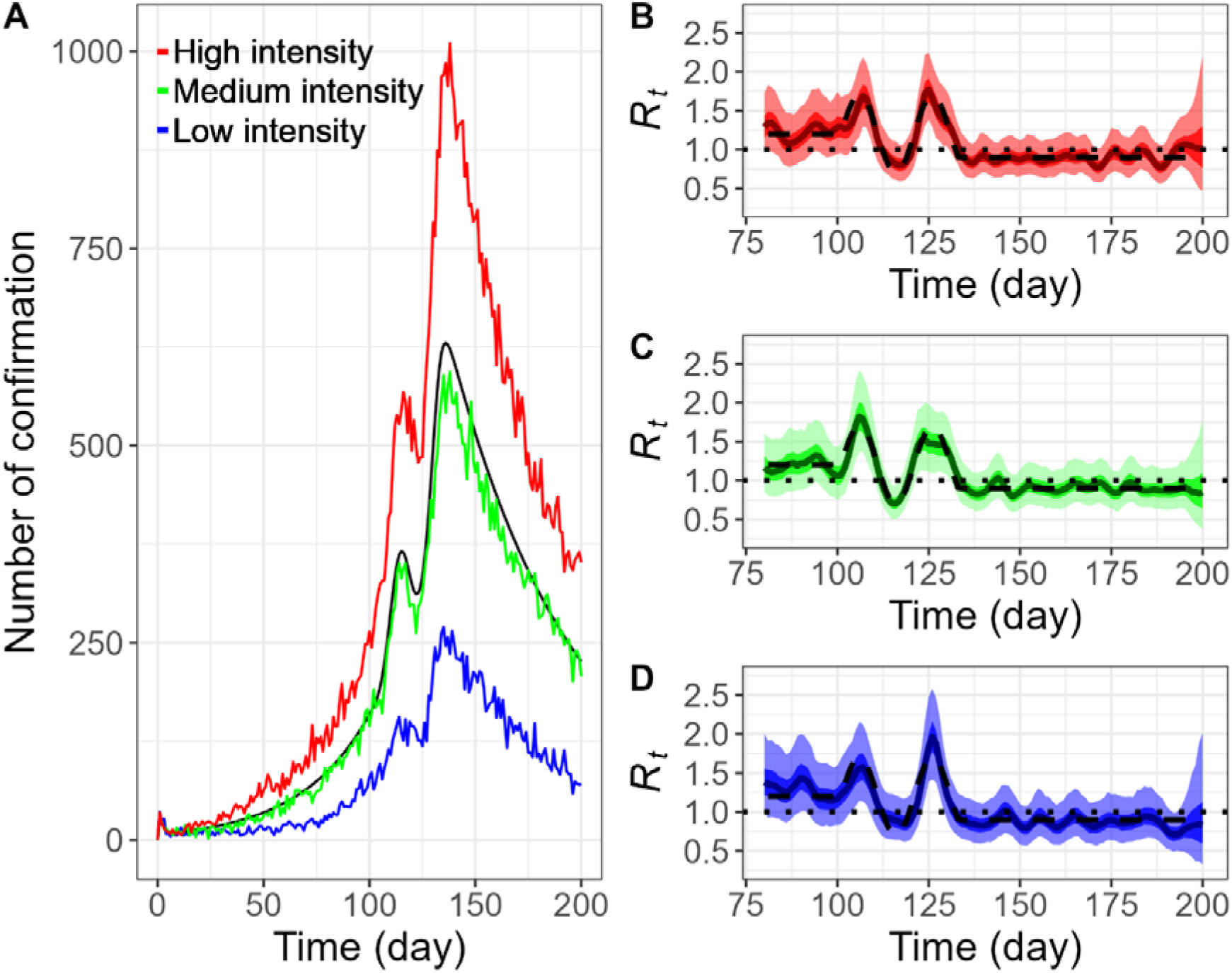
Daily confirmation time series and daily reproduction number, *R*_*t*_ inferred by particle filtering applied on stochastic SEPAIR models with*N*_*p*_=10,000 and *N*_*b*_=1,000. Three samples are chosen on a basis of low, medium, and high incidence, all with initial infection size of 100 (i.e.*I*_0_=100). **A**. Plots of *R*_*t*_daily confirmation time series for deterministic simulation (black); and stochastic simulation with low intensity (blue), medium intensity (green), and high intensity (red). **B**., **C**., **D**. Plots of *R*for low intensity (blue), medium intensity (green), and high intensity (red), respectively: Pre-defined *R*_*t*_(black dashed), reference line (black dotted), median of estimated *R*_*t*_(dark blue, green, red line resp.), interquartile range of estimated *R*_*t*_(blue, green, red shaded resp.), and middle 95% of estimated *R*_*t*_ (light blue, green, red shaded).

### 3.2. Estimation of *R*_*t*_based on a renewal equation method

#### 3.2.1. Inferring *R*_*t*_using confirmation times series based on a stochastic simulation observation model

Instantaneous *R*_*t*_based on a renewal equation implemented in EpiEstim also captures the true *R*_*t*_when the time series of infection was provided. However, the delay is observed when the time series of confirmation is directly used (see Fig 5), and this can be corrected with by using an appropriate information delay from infection to confirmation. Otherwise, the renewal equation-based strategy can capture overall shape of the pre-defined *R*_*t*_well. In our case, infection time series was estimated through deconvolution operations (Gostic et al., 2020), which involves adjustment of time points by 4 days backward in time. Such an adjustment is based on provided information of delay distribution, but causes early termination of estimation procedure, e.g., no estimates are provided in the last 4 days (Fig 6, Fig 7). Then we employed EpiEstim method with a serial interval (mean = 6.25 days and standard deviation = 4.14578 days), and the three time series of confirmed cases used in the earlier PF estimation. Each time series was preprocessed to obtain a moving average with lookback days of 7 (i.e., a week). This step reduces unnecessary fluctuations but, together with deconvolution, results in over-smoothed *R*_*t*_curves (Fig 6). In particular, Fig 7 illustrates under-estimation of the peaks on days around 107 and 125 by significant amounts for all three estimations. Furthermore, we examined an effect of misspecification of serial interval by taking a smaller mean of 5 days. Such a reduction amplifies the under-estimation of *R*_*t*_curves around the peaks though little effects on the rests (Fig 7). The shorter serial interval perhaps makes estimation less adaptive to rapidly changing transmission patterns.

**Fig. 5.**
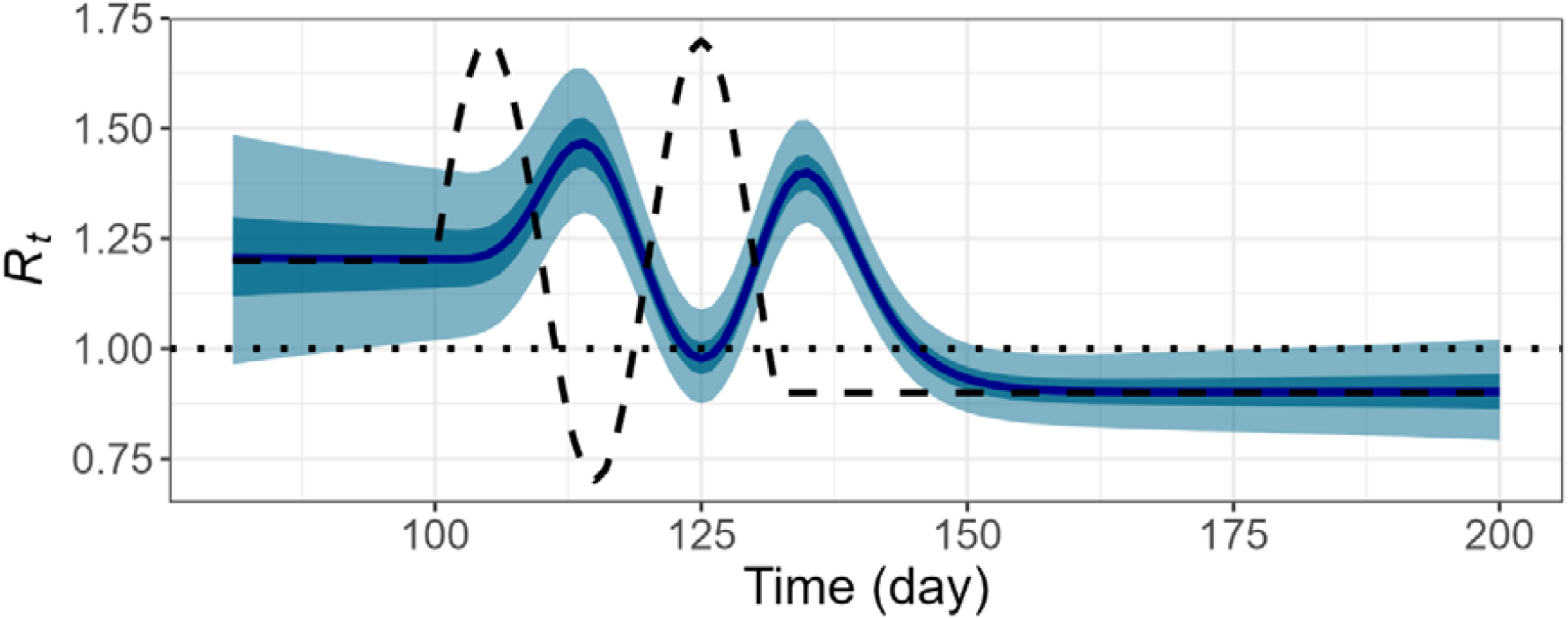
Daily reproduction number, *R*_*t*_, inferred by applying EpiEstim to the confirmation time series based on a deterministic model. Pre-defined *R*_*t*_(black dashed), reference line (black dotted), median of estimated *R*_*t*_(dark blue), interquartile range of estimated *R*_*t*_(dark cyan shaded), and middle 95% of estimated *R*_*t*_(light cyan shaded).

**Fig. 6.**
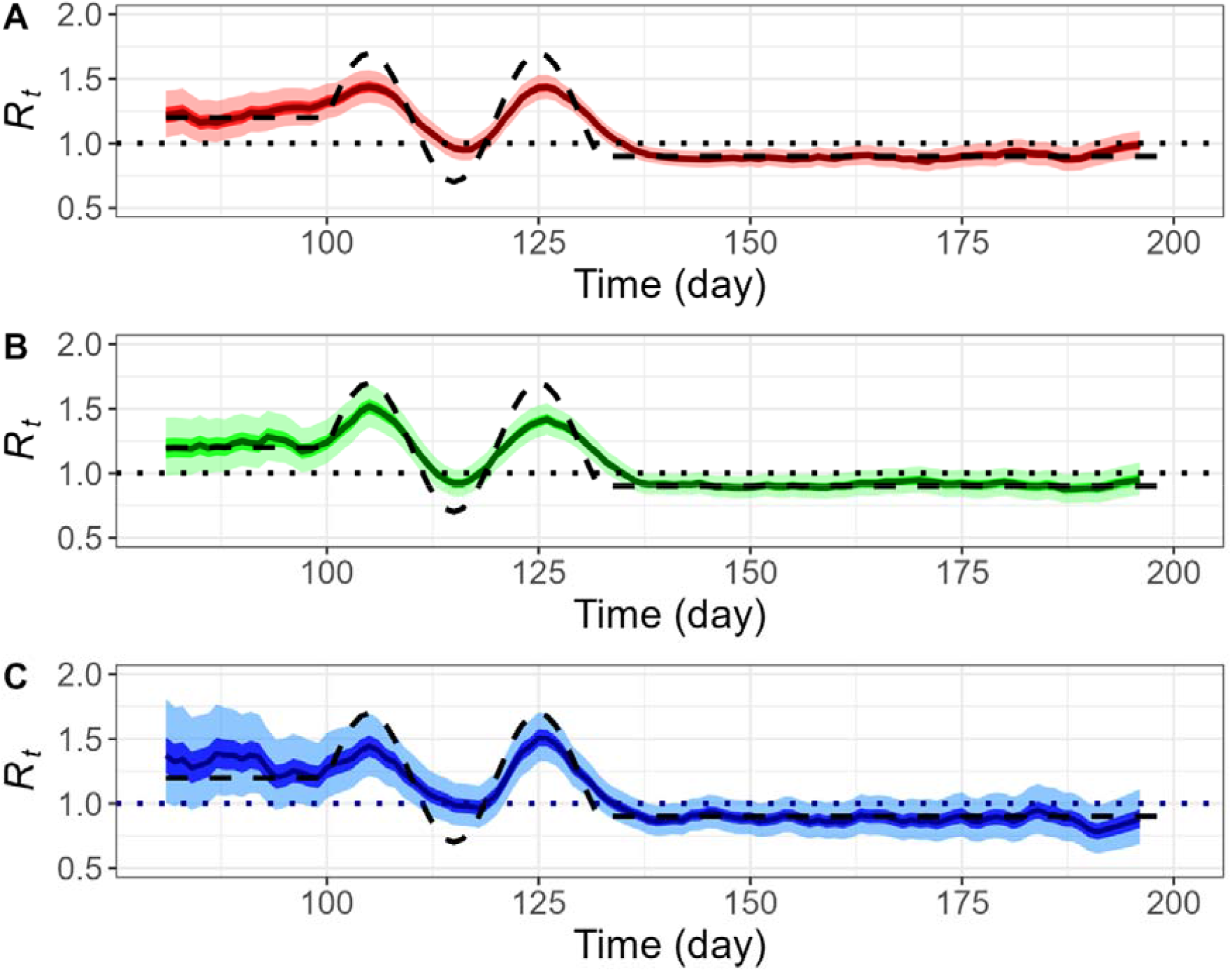
Daily reproduction number, *R*_*t*_ inferred by deconvolution followed by EpiEstim based on stochastic SEPIAR models with perfect observation. Three samples are chosen on a basis of low, medium, and high incidence, all with initial infection size of 100 (i.e.*I*_0_=100). **A**., **B**., **C**. Plots of *R*_*t*_for low intensity (blue), medium intensity (green), and high incidence (red), respectively: Pre-defined *R*_*t*_(black dashed), reference line (red dotted), median of estimated (dark blue, green, red line resp.), interquartile range of estimated *R*_*t*_(dark blue, green, red shaded resp.), and middle 95% of estimated *R*_*t*_(light blue, green, red shaded resp.).

**Fig. 7.**
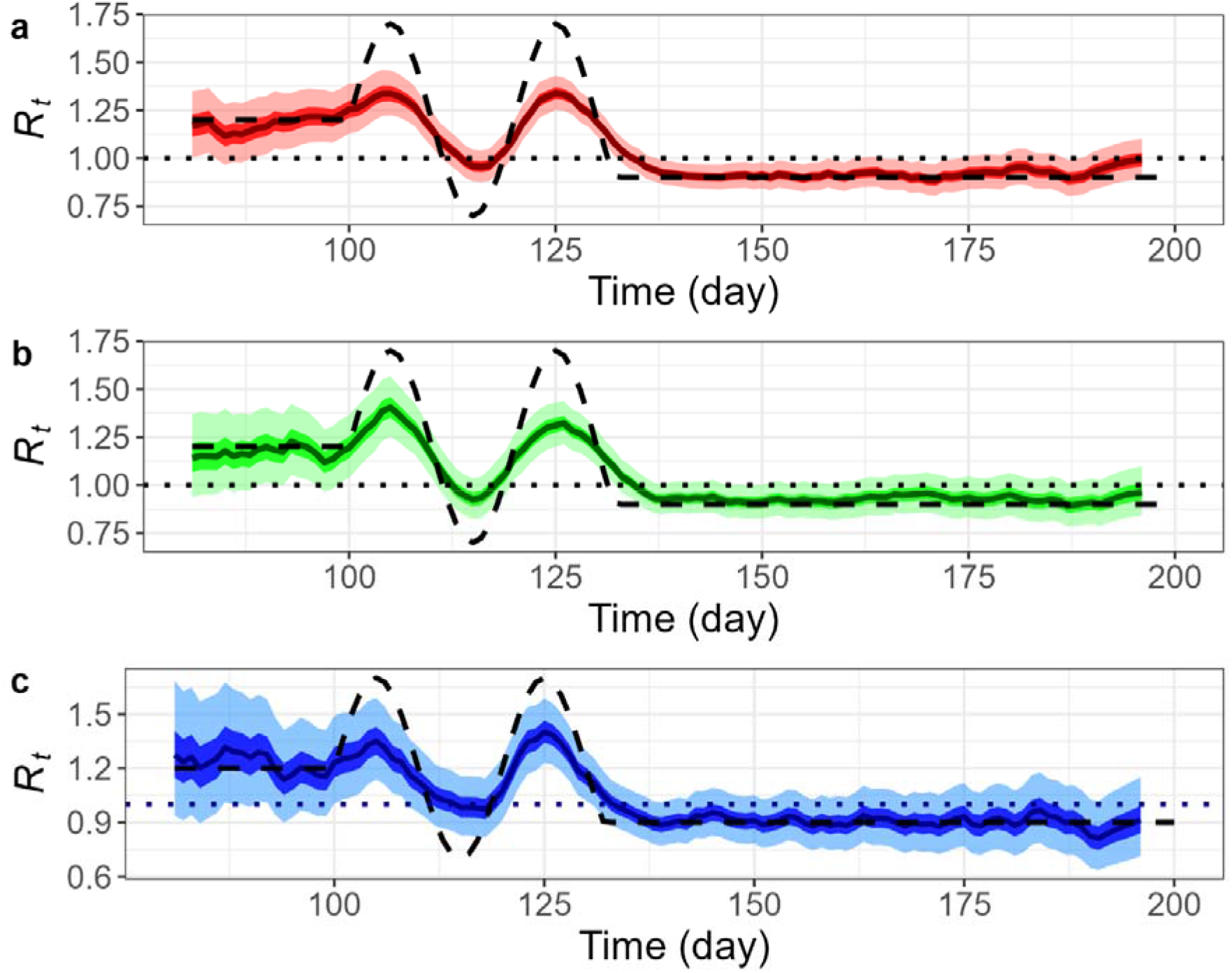
Daily reproduction number, *R*_*t*_ inferred by deconvolution followed by EpiEstim based on stochastic SEPIAR models with perfect observation and a misspecification of serial interval (mean = 5 days). Three samples are chosen on a basis of low, medium, and high incidence, all with initial infection size of 100 (i.e.*I*_0_=100). **A**., **B**., **C**. Plots of *R*_*t*_for low intensity (blue), medium intensity (green), and high intensity (red), respectively: Pre-defined*R*_*t*_(black dashed), reference line (red dotted), median of estimated *R*_*t*_(dark blue, green, red line resp.), interquartile range of estimated *R*_*t*_(dark blue, green, red shaded resp.), and middle 95% of estimated *R*_*t*_(light blue, green, red shaded resp.).

### 3.3. Performance comparison of two *R*_*t*_estimation methods

We calculated the RMSE score (Eq. 2) to assess the performance of the proposed PF method and the renewal equation-based method. The median of *R*_*t*_estimates based on the confirmation time series from the dataset v2 (the sample of medium incidence used in Sec 3.2.) was compared against to the pre-defined *R*_*t*_for 5 different periods (the simulation period from day 80 to day 120, and 4 different parts of it by epidemic characteristics) as indicated in Fig 7 and Table 2.

**Table 2.** RMSE Scores of inferred by two different methods for different periods.

|  | Particle Filter | Renewal Equation | Renewal Equation<br>(with misspecification effect) |
| --- | --- | --- | --- |
| **Period from day 80 to day 200** | 0.0983 | 0.0977 | 0.1211 |
| **Flat period** | 0.0512 | 0.0426 | 0.0396 |
| **Last 1 week of flat period** | 0.0119 | 0.0069 | 0.0098 |
| **Fluctuation period** | 0.1779 | 0.2250 | 0.2751 |
| **Last 1 week of fluctuation period\*** | 0.2495 | 0.4449 | 0.5130 |

\* NB. Simulation ended on the last day of the fluctuations (i.e., day = 132). The RMSE were calculated for every available value within an indicated period.

Both the methodologies showed similar performance for the general period of day 80 to day 200. The renewal equation-based strategy outperforms the PF slightly, when the true *R*_*t*_follows a steady progression (i.e., flat). It can be explained by the delay from infection to confirmation (i.e., mean delay = 7.5 days), which was addressed by deconvolution and moving average operations in prior to the EpiEstim operations. Another factor could be stochasticity contained in PF. Such an observation becomes clearer in the last 1 week of the simulation, where the RMSE of PF is noticeably greater than of the renewal equation (Table 2). In contrast, the PF outperforms the renewal equation for dynamic periods of epidemic transmissions (i.e., cycles of massive infections). It can be explained by the adaptive feature of PF, and over-smoothing caused by deconvolutions and moving average operations.

Particularly, the misspecification of a shorter serial interval led to worse performance. An additional simulation was conducted until the last day of the infection cycles (i.e., day 132) and the gap of RMSE scores amongst three experiments widened expectedly (Table 2, Figure 9). Note that the renewal equation-based estimations (Figures 9B, 9C) ended 4 days earlier than the PF estimation (Figure 9A) as the results of deconvolution.

**Fig. 8.**
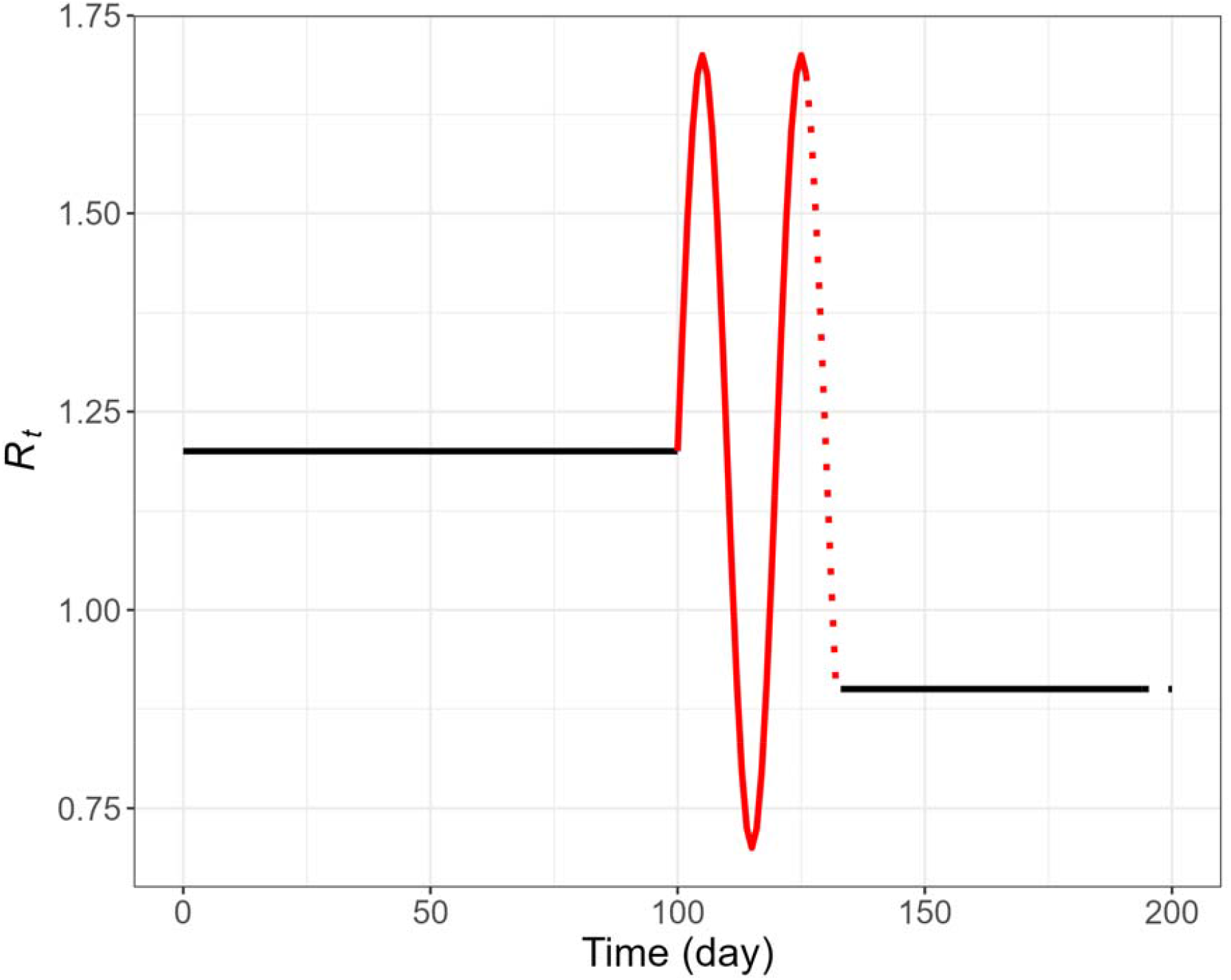
The curve of the pre-defined daily reproduction number, *R*_*t*_, indicated by 4 different periods: flat period (in black), last 1 week of the flat period (black dotted line), fluctuation period (in red), and last 1 week of the fluctuation period (red dotted line).

**Fig. 9.**
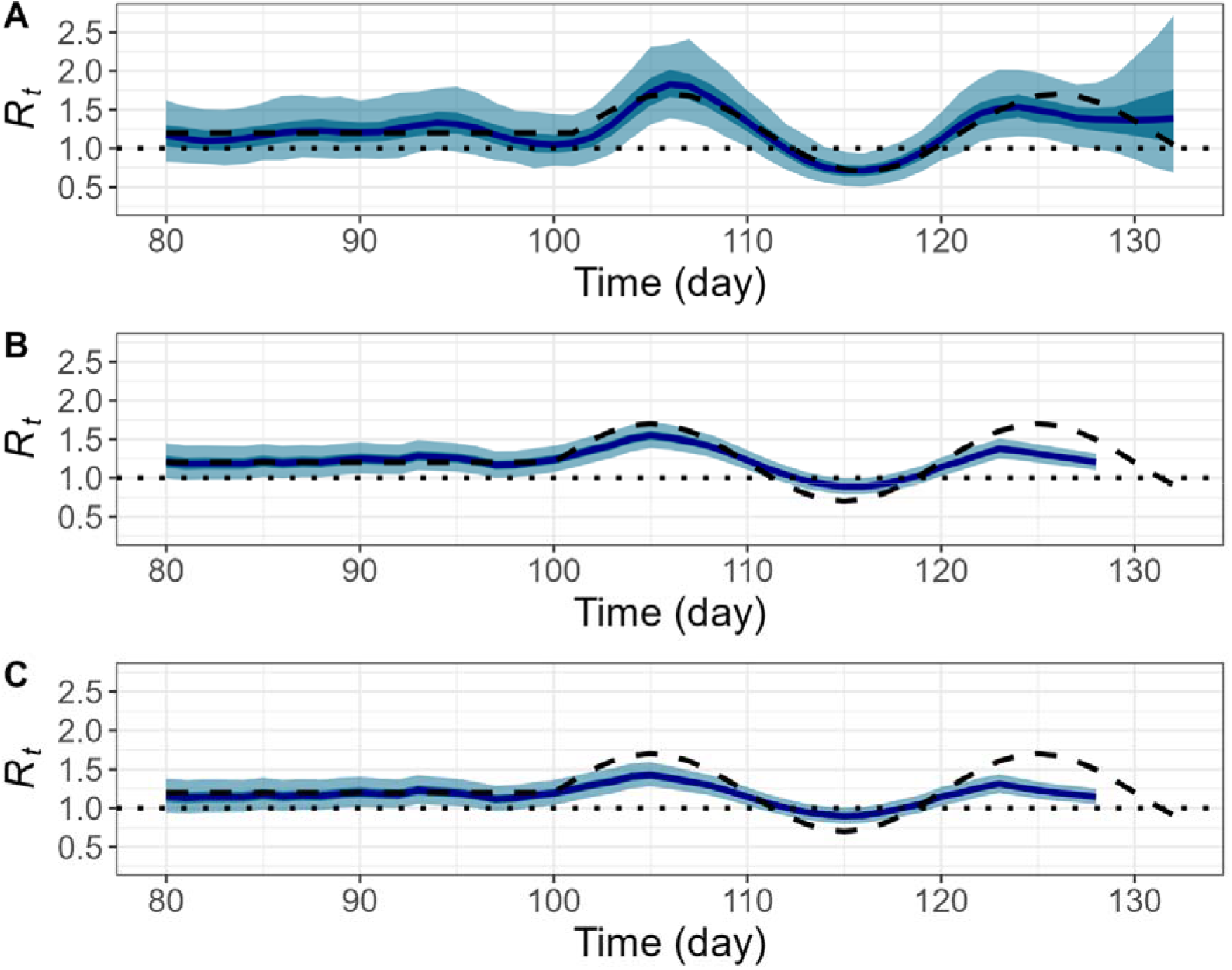
Plots of daily reproduction number, *R*_*t*_based on stochastic SEPIAR models with perfect observation (the medium incidence with initial infection size of 100 as in Sec. 3). Pre-defined*R*_*t*_ (black dashed), reference line (black dotted), median of estimated *R*_*t*_(dark blue), interquartile range of estimated (dark cyan shaded), and middle 95% of estimated *R*_*t*_(light cyan shaded). **A**.*R*_*t*_ inferred by particle filtering with *N*_*p*_=10,000and *N*_*b*_=1,000. **B**. *R*_*t*_inferred by deconvolution followed by EpiEstim. **C**.*R*_*t*_ inferred by deconvolution followed by EpiEstim, with a misspecification of serial interval (mean = 5 days).

## 4. Discussions

In this study, we proposed a particle filtering (PF) algorithm to estimate the effective reproduction number (*R*_*t*_) of an infectious disease using daily time series (e.g., of infection or confirmation) data. We compared the performance of the PF algorithm with a renewal equation-based *R*_*t*_estimation method, EpiEstim (Cori et al., 2013), and demonstrated the advantages in situations where transmission characteristics change rapidly or observation contains delays. We also introduced a framework of testing epidemic hypotheses and potential impact of intervention programs by constructing relevant scenarios based on an extended SEIR model (i.e., SEPIAR model).

Our approach is similar to the work (Kucharski et al., 2020) and we provide systematic analyses of the approach using simulated data that may represent various real-world scenarios. As with the Kalman filter approach (Arroyo-Marioli et al., 2021), our method can be used to extract “filtered” or “smoothed” estimate. Some recent studies adopt a Kalman filter approach to estimate *R*_*t*_based on a structural relationship between *R*_*t*_and a compartment model as developed in this paper (see Eq. 1). This owes to the notion of effective reproduction number introduced by Cori et al. (Cori et al., 2013). Arroyo-Marioli et al. (Arroyo-Marioli et al., 2020) used a Kalman filter and SIR equations by assuming a linear relation between *R*_*t*_and the growth rate of susceptible state (i.e., S), whereas Hasan et al. (Hasan et al., 2022) used an extended Kalman filter along with a SIRD model. The latter is seen as an improvement of the former as the nonlinear filter is employed and the underlying model is complicated with a notion of the case fatality rate. Our model takes the model of Hasan et al. further by addressing uncertainty in transmissions as pre-symptomatic and asymptomatic. We also used a nonlinear filter (i.e., particle filter) in view of that the more constituents in a compartment model would involve a greater degree of nonlinearity. Unlike the works above are validated by confirmation time series of different countries, our estimation model is evaluated by pre-determined scenarios of *R*_*t*_that capture realistic characteristics of epidemic transmissions. This provides a testing ground for both the estimation accuracy and misspecifications of the associated model.

The performance of the PF method decreases when the stochastic time series are given but still performs better than the commonly used EpiEstim method (Cori et al., 2013). EpiEstim method based on the renewal equation appears to present significant challenges before it is correctly applied to the confirmation time series of COVID-19 pandemic. One issue is to reliably infer infection time series from confirmation time series. The other issue is to estimate serial interval reliably from contact and symptom onset data, which may not be straightforward because of the pre-symptomatic transmission.

Our study has limitations. The SEPIAR model employed in this study makes a simplifying assumption that all infected people are observed or at least the detection rate stays constant over the simulation period. In reality, only a fraction of the infected people would be detected with time-varying probability of detection. To mitigate this unrealistic assumption, we used the stochastic time series in which daily incidence is larger than, similar to, and lower than the predictions by the deterministic model. The use of these stochastic time series may account for imperfect observation as well as stochasticity of the transmission process.

Second, we assumed that the parameters for the underlying model (SEPIAR) and EpiEstim are known. Such parameter values may be estimated but only with uncertainties or potential biases. This implies we are likely to overestimate the performance of our methods and we conducted a further experiment on the misspecification of serial interval with two different lengths (mean = 6.25 or 5 days). The robustness of PF-based predictions was confirmed by consistent trends of *R*_*t*_ -curves (see Fig 7, Fig 8). We believe that the superiority of the PF over the EpiEstim method would still be retained even if parameter values are unknown provided similar information. During the COVID-19 pandemic, we are confronted with confirmation time series which are based on limited testing of suspected cases, imperfect diagnosis, and significant delay from infection or symptom onset to confirmation. While our study accounts for several aspects of these realities such as delay from infection or symptom onset and partially imperfect diagnosis, additional aspects of realities need to be explored in developing a method to estimate *R*_*t*_.

## 5. Conclusions

Particle filtering method can be implemented in the context of an SEPIAR compartmental model and recover the true *R*_*t*_based on the simulated data, which mimic COVID-19 data. The model with filtered parameter values can serve as a framework to test hypotheses and explore potential impact of intervention programs during the COVID-19 pandemic.

## Data Availability

All data produced are available online at

https://github.com/kimfinale/pfilterCOVID

## Funding

This research was partly supported by Government-wide R&D Fund project for infectious di disease research (GFID), Republic of Korea (grant number: HG18C0088) and National Institute for Mathematical Sciences (NIMS) grant funded by the Korean Government (NIMS-B21910000)

## CRediT authorship contribution statement

**Jong-Hoon Kim**: Conceptualization, methodology, software, original draft preparation. **Yong Sul Won**: Original draft preparation, data curation, writing, and editing. **Woo-Sik Son**: writing, validation, editing. **Sunhwa Choi**: Writing, validation, editing.

## Acknowledgments

All authors acknowledge useful discussions with the members of the Research and Development on Integrated Surveillance System Development for Early Warning of Infectious Diseases of Korea.

## Notes

### Competing Interest Statement

The authors have declared no competing interest.

### Author Declarations

Source data were publicly available at Korea Disease Control Agency.

